# Genomic Studies in Blood and Marrow Transplantation (GS-BMT): An NCI-CIBMTR genomic medicine initiative in hematopoietic cell transplantation

**DOI:** 10.64898/2026.11.06.26367367

**Authors:** Shahinaz M. Gadalla, Namhwa Lee, Xueyao Wu, Filip Pirsl, Gabrielle Schmidt, Maryam Rafati, Amy Hutchinson, Aurelie Vogt, Belynda Hicks, Jia Liu, Jiahui Wang, Shilpa Gaddam, Shengchao Li, Wael Saber, Stephen Chanock, Peter Kraft, Sharon A Savage, Hyokyoung G. Hong, Stephen R Spellman

## Abstract

**Background:** Genomic studies in hematopoietic cell transplantation (HCT) are limited by sample sizes and targeted approaches.

**Methods:** The Genomic Studies in Blood and Marrow Transplantation (GS-BMT) project is a collaboration between the National Cancer Institute (NCI) and the CIBMTR® designed to investigate recipient and donor determinants of HCT outcomes. We are generating genome-wide SNP-array genotyping for all samples, and sequencing and methylation profiling for selected subsets. We report study design and cohort characteristics and present two proof-of-concept analyses: comparison of conventional and machine learning prediction of overall survival after HCT in 9,262 patients with acute myeloid leukemia (AML), and assessment of inherited susceptibility to clonal hematopoiesis of indeterminate potential (CHIP) among 19,750 HCT donors using polygenic scores (PGS).

**Results:** GS-BMT includes 30,718 donor–recipient pairs who received allogeneic HCT in the United States between 2000 and 2019. In AML, clinical variables alone provided modest prediction for post-transplant survival, with similar performance for Cox proportional hazards, random survival forest, and deep neural network survival models (C-index=0.63, 0.62, and 0.62, respectively). In donors, CHIP-PGS did not differ by age (p=0.76) but was modestly lower in unrelated *vs*. related donors (−0.06 standard deviation [SD]; p=0.002). In an illustrative analysis assuming risk of event increases by 50% or 80% per PGS-SD, the top 5.2% and 12.3% of the donors, respectively, would have at least twice the risk of a donor with the average PGS.

**Conclusions:** GS-BMT establishes a large genomic platform aiming to improve individualized risk prediction, donor selection, and precision medicine in HCT.

## Introduction

Allogeneic hematopoietic cell transplant (HCT) is a well-recognized curative therapy for several life-threatening hematologic diseases, with >8,000 recipients annually in the United States.^1^ Acute leukemias, myelodysplastic syndrome (MDS), and myeloproliferative neoplasms (MPN) combined account for approximately two-thirds of the cases.^1,2^ HCT methods have advanced over the last two decades leading to significant survival improvements (3-year probability of overall survival= 45.3% in 2002-2006 *versus* 62.1% in 2017-2022).^3^ Some of the recent advances include the introduction of post-transplant cyclophosphamide for graft-*versus*-host disease (G*v*HD) prophylaxis that allowed for expanding donor selection to haplo-identical related and human leukocyte antigen (HLA) mismatched unrelated donors, the development of new immunomodulatory agents to treat severe chronic G*v*HD, and the development of new conditioning regimens that decreased organ toxicity and expanded transplant eligibility to older patients.^4,5,6^

The significant role of genomics in HCT was recognized from its inception with a focus on immunogenetics particularly for HLA genes. HCT genomic studies beyond HLA are still sparse, with most findings derived from relatively small studies (∼60 to 1000 patients; reviewed in^7–9^) using candidate genes or similarly targeted approaches. Examples of these studies include associations between donor *IFNL4* genotype and patient overall survival (OS) or transplant-related mortality (TRM), and *CTLA-4* and disease-free survival (DFS) and OS.^10–12^ DISCOVeRY-BMT was the first genome-wide association study (GWAS) in HCT, including 3,419 patients with acute leukemia or MDS, and focused on one-year post-HCT mortality. Although limited by its relatively small sample size for GWAS and heterogeneity in disease- and transplant-related factors, DISCOVeRY-BMT showed several promising findings.^13,14^

The Genomic Studies in Blood and Marrow Transplantation (GS-BMT) project is a collaboration between the National Cancer Institute (NCI) and the CIBMTR® (Center for International Blood and Marrow Transplant Research), representing the largest HCT genomic project to-date. GS-BMT is designed to allow for the inclusion of both malignant and non-malignant diseases, all ages and ethnic groups, and different treatment regimens. The project includes SNP-array genotyping for all recipient-donor pairs as well as sequencing and methylation array for selected cohort subsets. This report describes the GS-BMT methodology and population characteristics, evaluates different predictive tools of HCT survival in the GS-BMT AML cohort using clinical information and presents an example of the effective use of these genomic data in donor selection focusing on susceptibility to clonal hematopoiesis of indeterminate potential (CHIP). Current donor selection practices prioritize young donors (typically <30 years of age), which reduces the likelihood of prevalent CHIP, but likely does not control for its underlying genetic predisposition.

### GS-BMT Design and Methodology

#### Source of Data and Blood Samples

The GS-BMT uses blood samples and clinical data collected by the CIBMTR® (a research collaboration between the Medical College of Wisconsin and the NMDP (formerly National Marrow Donor Program and Be The Match). CIBMTR compiles extensive clinical information and HCT outcome data and houses a large biorepository of donor (pre-collection) and recipient (pre-conditioning regimen) biospecimens for unrelated donor transplants performed in the U.S. since 1987 and for related donor HCT since 2006; details are available at https://cibmtr.org. Data collected by the CIBMTR are audited to ensure completeness and high quality and represent the real-world clinical practice experience (https://cibmtr.org/CIBMTR/Data-Operations/Data-Operations-Compliance/Audit-Program).

#### GS-BMT Primary Aims & End Points

The study aims to 1) identify recipient and donor germline genetic variation and somatic alterations associated with HCT outcomes; 2) develop individualized risk calculators for post-HCT overall and event-free survival by incorporating key genomic and clinical factors; 3) characterize the germline genetic etiology of selected hematological malignancies (both common and rare risk variants); and 4) build and maintain a robust genomic data resource available for use by the HCT scientific community.

The study primary end points are all-cause and cause-specific mortality, and disease relapse. Secondary end points include: 1) G*v*HD-free, relapse-free survival, defined as survival without grade III-IV acute G*v*HD, chronic G*v*HD requiring systemic therapy, or relapse; and 2) risk of subsequent malignancies

#### Eligibility Criteria

HCT recipients were eligible to be included in the study if they: 1) received HCT at a U.S. center between 2000-2019; and 2) had available clinical data and pre-HCT blood sample for both recipient and corresponding donor. The study excluded cord transplants, diseases for which allogeneic HCT is not a standard of care (*e.g.,* lymphoma), and rare HCT indications (<300 cases during the study period). A total of 34,058 recipient-donor pairs met the study inclusion criteria; of them 30,718 consented to research-based genetic testing including deposition of genetic data in controlled access public databases and were included in this study. All participants or their legal guardian provided written informed consent for the research use of their samples and clinical data.

#### DNA Extraction, Genotyping, and Imputation

The laboratory workflow was performed at the NCI Cancer Genomics Research Laboratory (CGR; https://dceg.cancer.gov/about/organization/cgr). DNA was extracted from frozen whole blood or peripheral mononuclear cells using an automated high-throughput bead-based method (ThermoFisher; KingFisher). Samples were genotyped on the customized Illumina Infinium Global Screening Array version 3.0 (GSA-V3), which interrogates approximately 743,000 markers with enrichment for low-frequency variants and multi-ethnic coverage. Genotypes were called using the standard Illumina array-analysis workflow with a study-specific cluster file and aligned to genome build GRCh38.

Genotyping quality control (QC) included sample- and variant-level quality filters, sample identity verification (contamination, sex, technical replicates), and population structure characterization (ancestry inference, relatedness, genetic principal components [PCs]). Imputation was carried out against the TOPMed r3 reference panel. Full details of genotyping, imputation, and QC metrics are provided in the Supplemental Methods.

#### Germline Risk of Clonal Hematopoiesis of indeterminate potential (CHIP) in HCT Donors

To characterize the germline susceptibility to CHIP among HCT donors, we constructed a CHIP polygenic score (PGS) using genetic variants and weights from a recent large-scale GWAS.^15^ Of 57 reported index variants, 51 were available and met imputation quality thresholds in our imputed data and were included in the score (Supplementary Table S1). PGS were calculated in PLINK2 using --score without mean imputation. Weighted allele sums were combined across chromosomes, divided by the total number of alleles included for each donor. Individual raw PGS were standardized to z-scores across the donor cohort, where a z-score of zero represents the cohort mean.

We then assessed standardized PGS by donor age (<35 vs ≥35 years) and donor type (related *vs* unrelated to their corresponding HCT recipient with a hematological malignancy) using linear regression, adjusting for the top eight genetic principal components.

To illustrate the potential clinical relevance of inherited CHIP susceptibility, we calculated the proportion of donors whose standardized PGS would correspond to at least twice the risk of a hypothetical adverse outcome compared to a donor with the mean PGS. This population proportion was determined under three separate modeling assumptions, in which each additional standard deviation (SD) in PGS was associated with a 20%, 50%, or 80% higher risk of the hypothetical adverse outcome.

## Results

### Characteristics of the GS-BMT population

GS-BMT includes 30,718 unique HCT recipients and 30,656 unique donors (55 of whom served as a donor for two recipients; seven donors withdrew consent and were excluded) from 238 transplant centers. Median recipient age at transplant was 50 years (range <1-83), while median donor age was 30 years (range <1-80). The most common indications for HCT were acute myeloid leukemia (AML; 46%) followed by MDS or MPN (22%) and acute lymphoblastic leukemia (ALL; 18%). Donors were predominantly unrelated (83%) and mostly 8/8 HLA-matched (76%; considering high resolution at HLA-A, -B, -C and -DRB1) to recipients. Most recipients received donor peripheral blood stem cells (72%; Table 1)

**Table 1:**
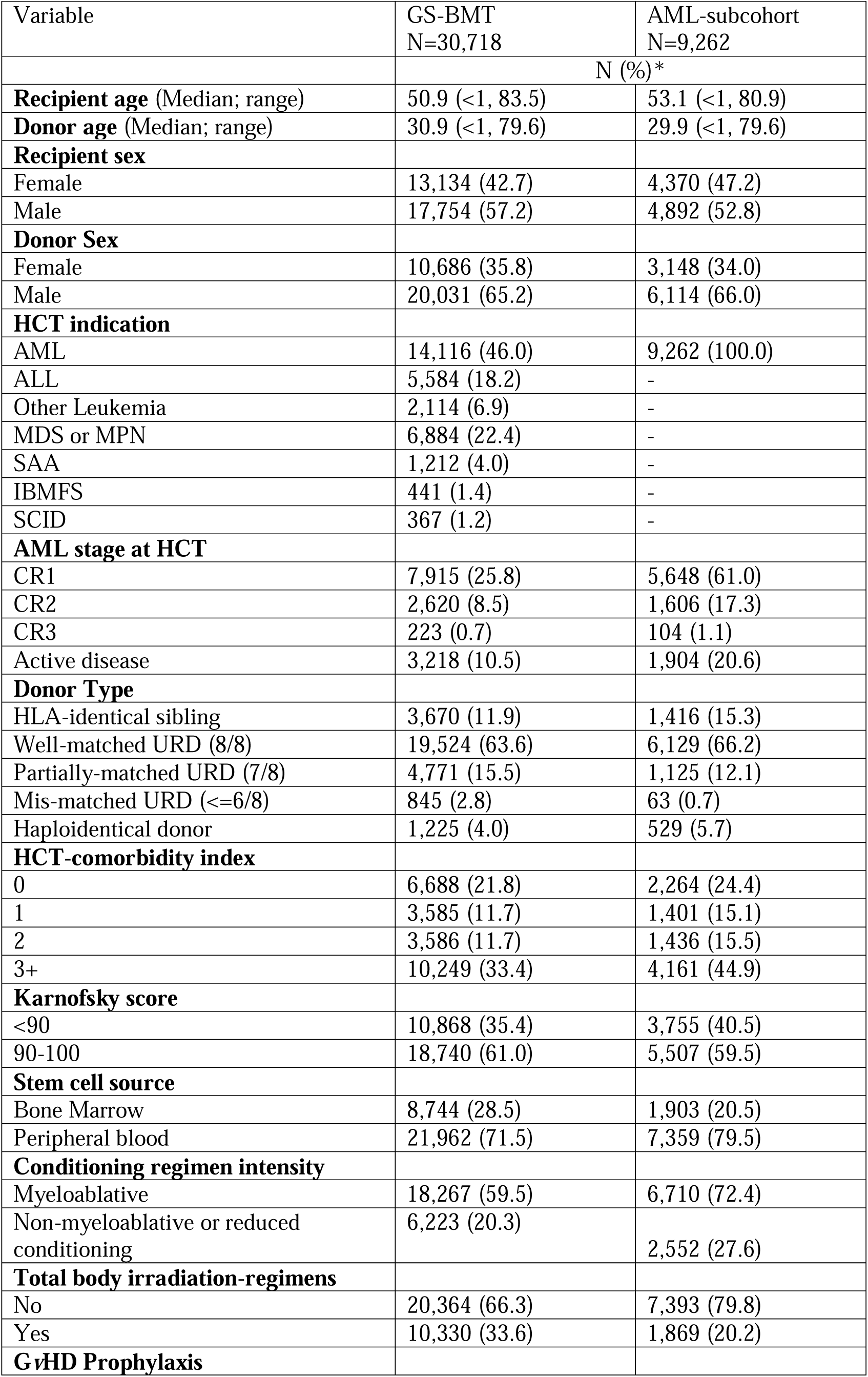
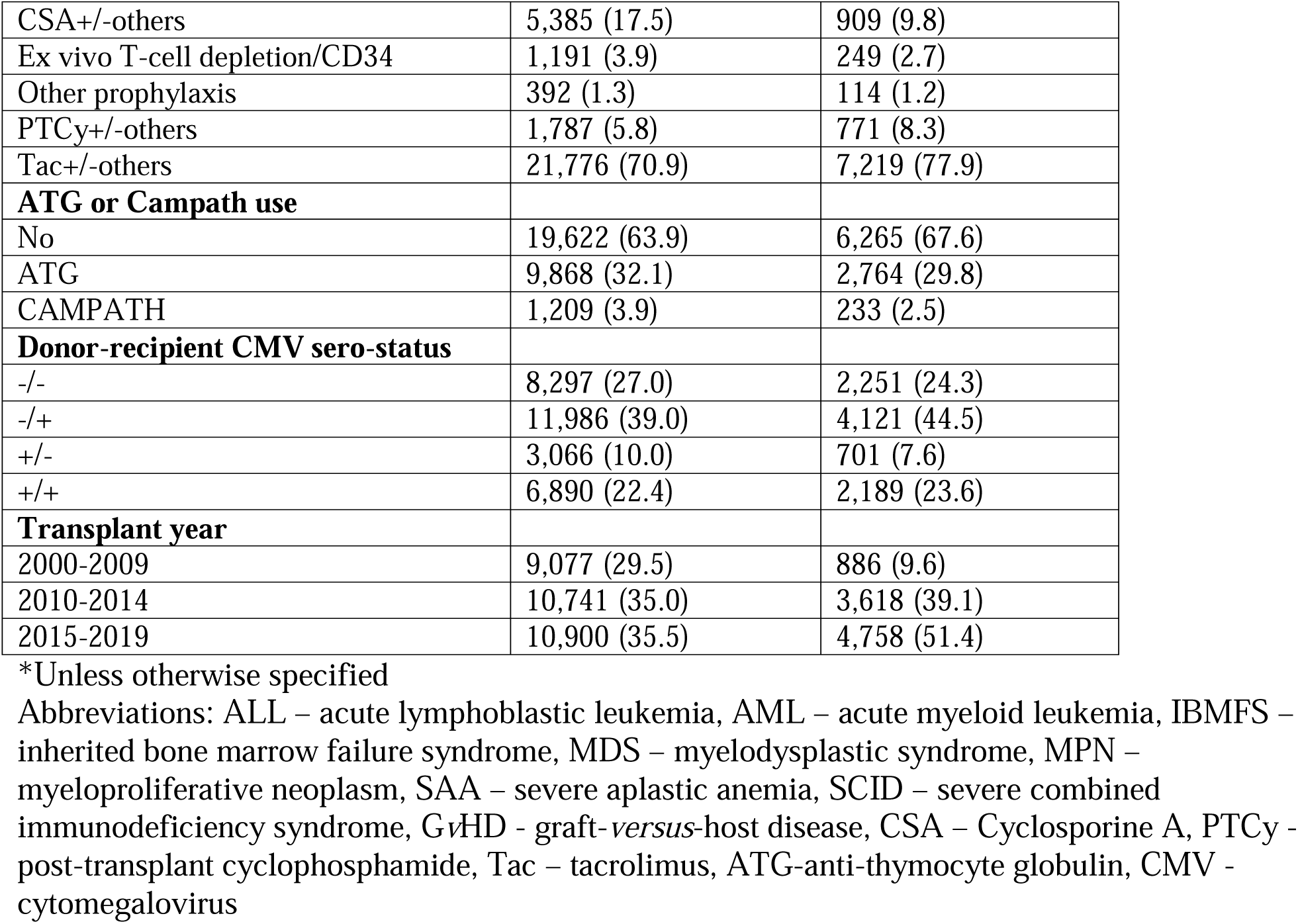
Demographics, clinical and transplant factors for GS-BMT participants.

| Variable | GS-BMT<br>N=30,718 | AML-subcohort<br>N=9,262 |
|---|---|---|
|  | N (%)* |  |
| **Recipient age** (Median; range) | 50.9 (<1, 83.5) | 53.1 (<1, 80.9) |
| **Donor age** (Median; range) | 30.9 (<1, 79.6) | 29.9 (<1, 79.6) |
| **Recipient sex** |  |  |
| Female | 13,134 (42.7) | 4,370 (47.2) |
| Male | 17,754 (57.2) | 4,892 (52.8) |
| **Donor Sex** |  |  |
| Female | 10,686 (35.8) | 3,148 (34.0) |
| Male | 20,031 (65.2) | 6,114 (66.0) |
| **HCT indication** |  |  |
| AML | 14,116 (46.0) | 9,262 (100.0) |
| ALL | 5,584 (18.2) | - |
| Other Leukemia | 2,114 (6.9) | - |
| MDS or MPN | 6,884 (22.4) | - |
| SAA | 1,212 (4.0) | - |
| IBMFS | 441 (1.4) | - |
| SCID | 367 (1.2) | - |
| **AML stage at HCT** |  |  |
| CR1 | 7,915 (25.8) | 5,648 (61.0) |
| CR2 | 2,620 (8.5) | 1,606 (17.3) |
| CR3 | 223 (0.7) | 104 (1.1) |
| Active disease | 3,218 (10.5) | 1,904 (20.6) |
| **Donor Type** |  |  |
| HLA-identical sibling | 3,670 (11.9) | 1,416 (15.3) |
| Well-matched URD (8/8) | 19,524 (63.6) | 6,129 (66.2) |
| Partially-matched URD (7/8) | 4,771 (15.5) | 1,125 (12.1) |
| Mis-matched URD (<=6/8) | 845 (2.8) | 63 (0.7) |
| Haploidentical donor | 1,225 (4.0) | 529 (5.7) |
| **HCT-comorbidity index** |  |  |
| 0 | 6,688 (21.8) | 2,264 (24.4) |
| 1 | 3,585 (11.7) | 1,401 (15.1) |
| 2 | 3,586 (11.7) | 1,436 (15.5) |
| 3+ | 10,249 (33.4) | 4,161 (44.9) |
| **Karnofsky score** |  |  |
| <90 | 10,868 (35.4) | 3,755 (40.5) |
| 90-100 | 18,740 (61.0) | 5,507 (59.5) |
| **Stem cell source** |  |  |
| Bone Marrow | 8,744 (28.5) | 1,903 (20.5) |
| Peripheral blood | 21,962 (71.5) | 7,359 (79.5) |
| **Conditioning regimen intensity** |  |  |
| Myeloablative | 18,267 (59.5) | 6,710 (72.4) |
| Non-myeloablative or reduced conditioning | 6,223 (20.3) | 2,552 (27.6) |
| **Total body irradiation-regimens** |  |  |
| No | 20,364 (66.3) | 7,393 (79.8) |
| Yes | 10,330 (33.6) | 1,869 (20.2) |
| **G*v*HD Prophylaxis** |  |  |
| CSA+/-others | 5,385 (17.5) | 909 (9.8) |
| Ex vivo T-cell depletion/CD34 | 1,191 (3.9) | 249 (2.7) |
| Other prophylaxis | 392 (1.3) | 114 (1.2) |
| PTCy+/-others | 1,787 (5.8) | 771 (8.3) |
| Tac+/-others | 21,776 (70.9) | 7,219 (77.9) |
| **ATG or Campath use** |  |  |
| No | 19,622 (63.9) | 6,265 (67.6) |
| ATG | 9,868 (32.1) | 2,764 (29.8) |
| CAMPATH | 1,209 (3.9) | 233 (2.5) |
| **Donor-recipient CMV sero-status** |  |  |
| -/- | 8,297 (27.0) | 2,251 (24.3) |
| -/+ | 11,986 (39.0) | 4,121 (44.5) |
| +/- | 3,066 (10.0) | 701 (7.6) |
| +/+ | 6,890 (22.4) | 2,189 (23.6) |
| **Transplant year** |  |  |
| 2000-2009 | 9,077 (29.5) | 886 (9.6) |
| 2010-2014 | 10,741 (35.0) | 3,618 (39.1) |
| 2015-2019 | 10,900 (35.5) | 4,758 (51.4) |

\*Unless otherwise specified
Abbreviations: ALL – acute lymphoblastic leukemia, AML – acute myeloid leukemia, IBMFS – inherited bone marrow failure syndrome, MDS – myelodysplastic syndrome, MPN – myeloproliferative neoplasm, SAA – severe aplastic anemia, SCID – severe combined immunodeficiency syndrome, G*v*HD - graft-*versus*-host disease, CSA – Cyclosporine A, PTCy - post-transplant cyclophosphamide, Tac – tacrolimus, ATG-anti-thymocyte globulin, CMV - cytomegalovirus

### Prediction Tools for Clinical Factors Impacting HCT Survival for Patients with AML

This analysis included the GS-BMT AML subcohort for whom clinical data on 15 *a priori* clinical factors were complete (N= 9,262; Table 1). We assessed prediction of overall survival (OS) by comparing a conventional multivariable Cox proportional hazards model with two machine learning (ML) approaches, a random survival forest (RSF) and a deep neural network survival model (survival DNN). See Supplemental Methods for details.

Results from multivariable Cox model are presented in Figure 1A & Supplementary Table S2. AML disease stage at HCT was the strongest predictor of OS with mortality risk increasing progressively from 1^st^ complete remission (CR1, reference) to active disease (hazard ratio, HR=1.1, p=0.01 for CR2; and 1.45, p=0.002 for CR3+; and 1.93, p=<0.001 for active disease). Other factors independently associated with OS included donor type with HLA identical sibling donors superior to matched unrelated or haplo-identical and alternative mismatched or partially matched donors, G*v*HD prophylaxis with cyclosporine-based regimens inferior to others, and cytomegalovirus (CMV) status with sero-negative donors for sero-positive recipients associated with mortality risk. In contrast, conditioning intensity, graft source, and ATG/Campath use were not significantly associated with post-HCT survival. The proportional hazards assumption was not met for several covariates (Supplementary Table S3), indicating that the effects of these variables on OS may vary over follow-up time.

**Figure 1:**
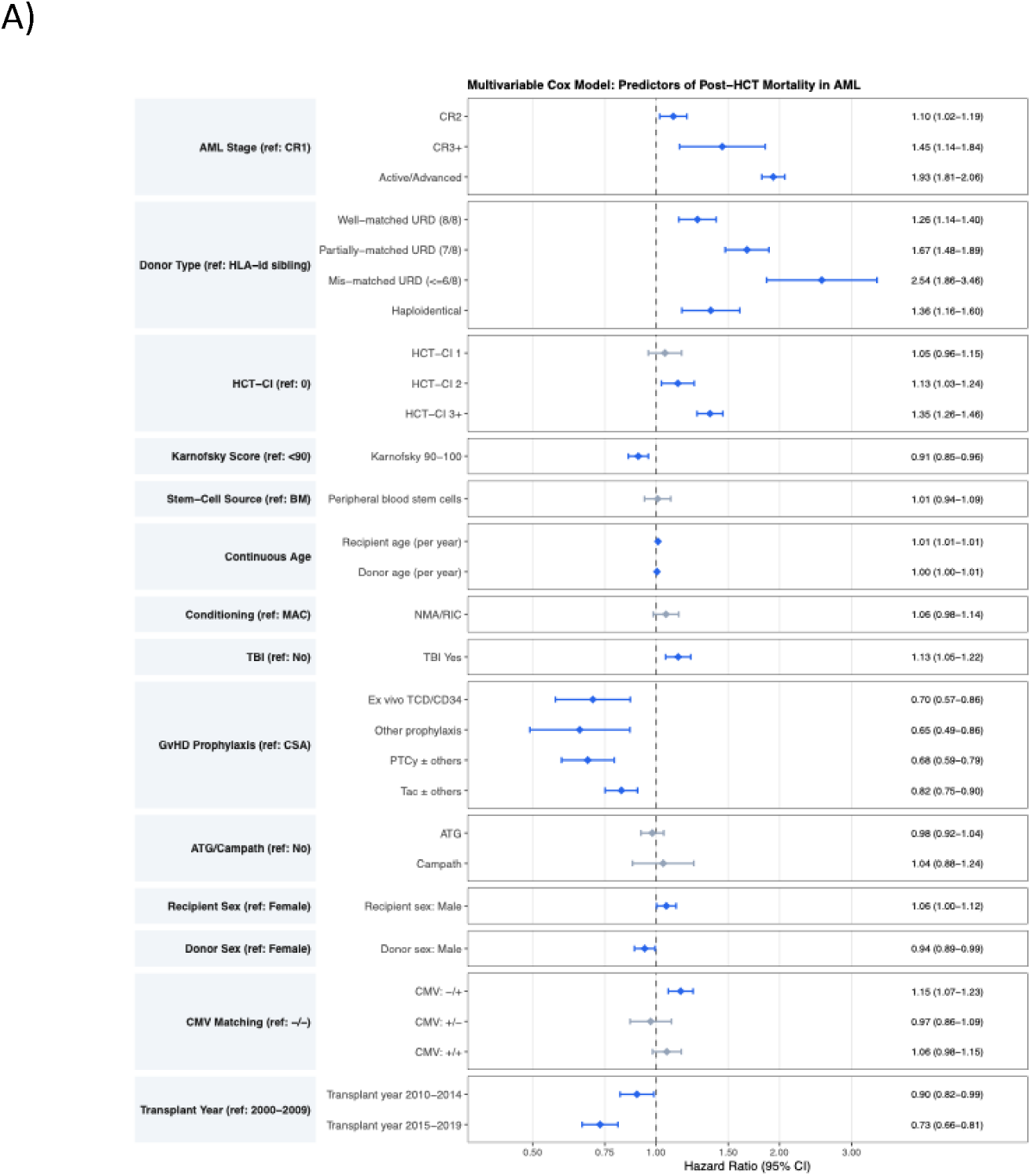
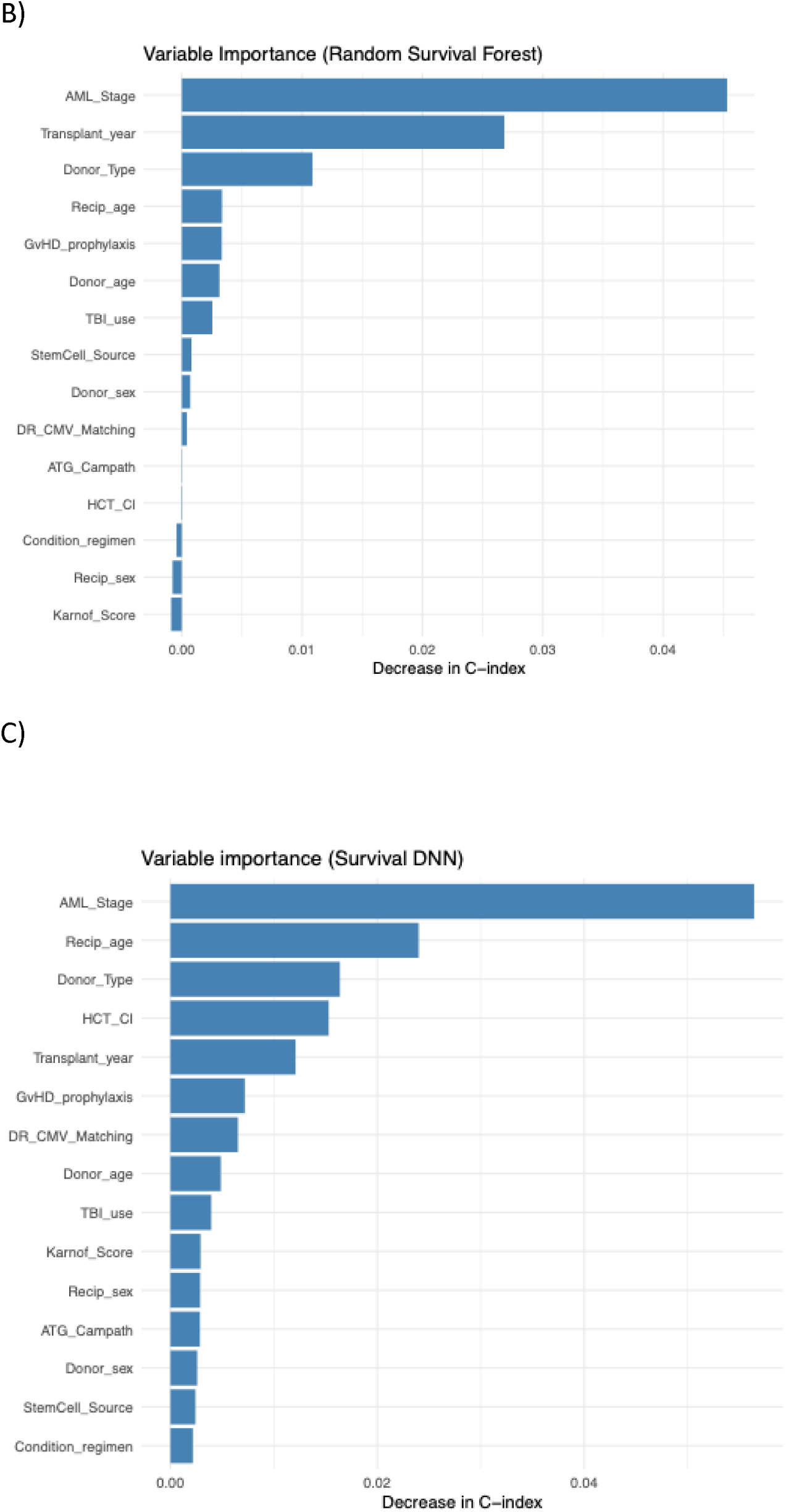
Clinical predictors of post-HCT overall survival identified using Cox proportional hazards and machine learning models. A) Multivariable Cox proportional hazards model for post-HCT overall survival in patients with AML. B) Variable importance from the random survival forest model for predicting post-HCT overall survival. C) Variable importance from the deep neural network survival model for predicting post-HCT overall survival.

In the machine-learning method, the RSF analysis also identified AML stage at HCT as the most important variable for predicting survival, followed by year of HCT, donor type, recipient age, and G*v*HD prophylaxis (Figure 1B). The survival DNN identified the same set identified by the RSF model, with the addition of the HCT comorbidity index (HCT-CI) (Figure 1C).

Classifying patients into three risk groups based on model-specific tertiles of risk scores showed clear separation of Kaplan–Meier curves (log-rank p < 0.001) with similar risk stratification across all three models (Figure 2). Model-specific continuous risk scores were highly correlated across the three approaches, with pairwise Spearman correlation coefficients ranging from 0.90 to 0.96 (Supplementary Table S4). Consistent with this finding, classification of patients into low-, intermediate-, and high-risk groups showed substantial agreement across models (Supplementary Figure S1). Risk discrimination was similar across models, with C-index (0.63 for the Cox model, 0.62 for the RSF, and 0.62 for the survival DNN). All three models demonstrated lower prediction error than the Kaplan–Meier null model, indicating improved predicted accuracy over a model without covariate information. However, overall predictive performance was nearly identical across approaches, as reflected by similar integrated Brier scores (IBS: 0.214, 0.213, and 0.214, respectively) and overlapping time-dependent Brier score curves (Supplementary Figure S2).

**Figure 2:**
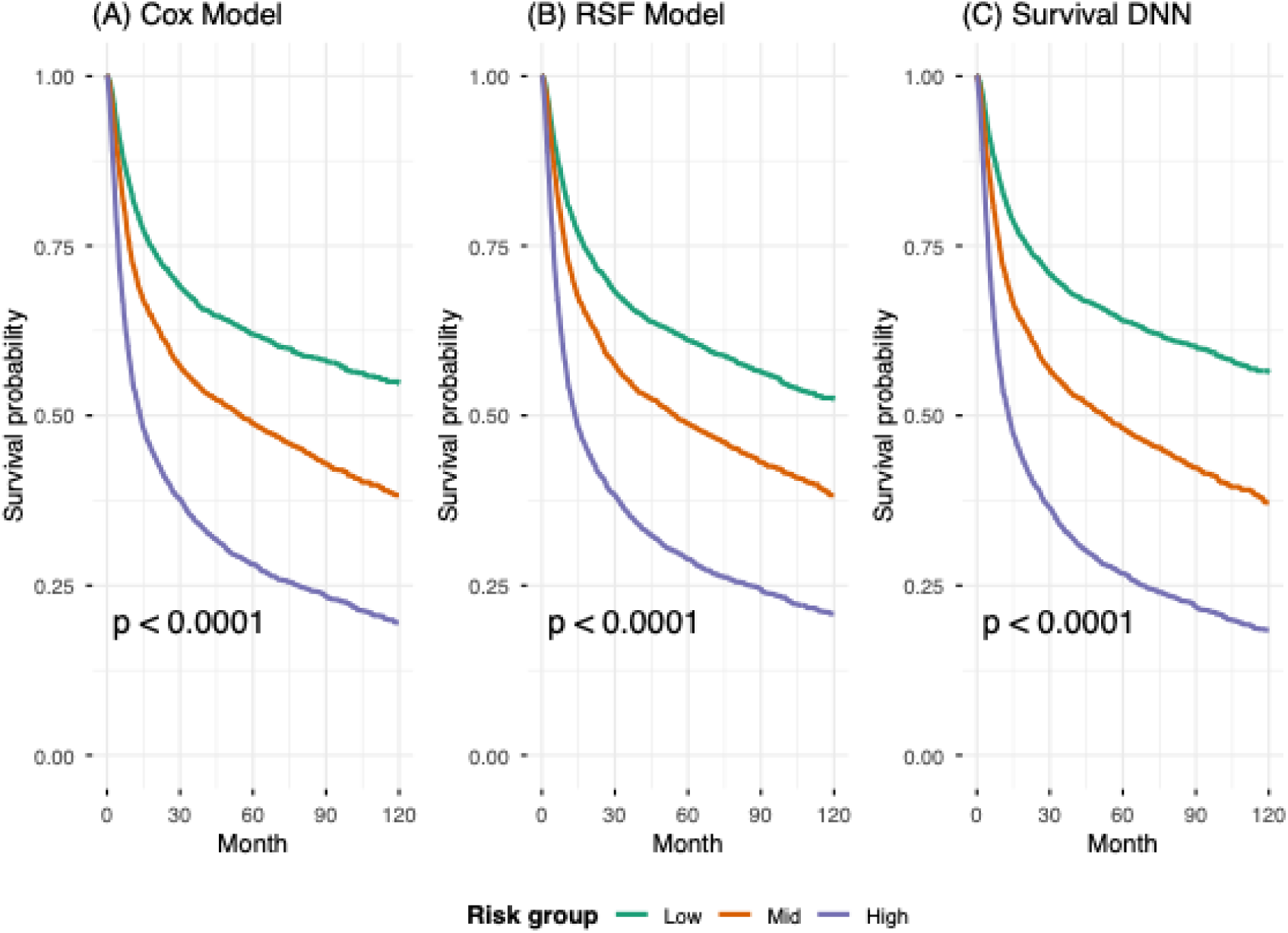
Kaplan-Meier curves by model-predicted risk group for post-HCT overall survival in patients with AML. Patients were stratified into low-, intermediate-, and high-risk groups according to tertiles of the model-specific risk scores generated by the (A) Cox proportional hazards model, (B) random survival forest (RSF), and (C) deep neural network survival model (survival DNN). Differences among risk groups were assessed using the log-rank test.

### Genotyping Performance and Genetic Ancestry

The first completed genotyping stage of the GS-BMT cohort successfully scanned 41,192 samples from recipient–donor pairs of patients with hematological malignancies, with a mean sample call rate of 98.7%. Imputed genotype data across 1–22 autosomes and the X chromosome were available for 39,554 unique subjects at 231,934,758 variants, including 19,804 recipients (10,939 AML, 4,068 ALL, and 4,797 MDS or MPN) and 19,750 donors. The imputed cohort was predominantly of European genetic ancestry (82.5%), followed by Latin American (9.5%) and African American (3.9%), with the remaining distributed across East Asian, Asian–Pacific Islander, South Asian, African, and other genetic ancestry groups (see Supplementary Methods for details on reference samples and algorithm used to define genetic ancestry groups). Genetic principal components and genetic ancestry coordinates were comparably distributed between recipients and donors (Supplementary Figure S3).

### Germline Risk of Clonal Hematopoiesis of Indeterminate potential (CHIP) in HCT Donors

Figure 3A shows the distribution of standardized CHIP PGSs among 19,750 HCT donors, with higher scores indicating greater inherited risk of CHIP

**Figure 3:**
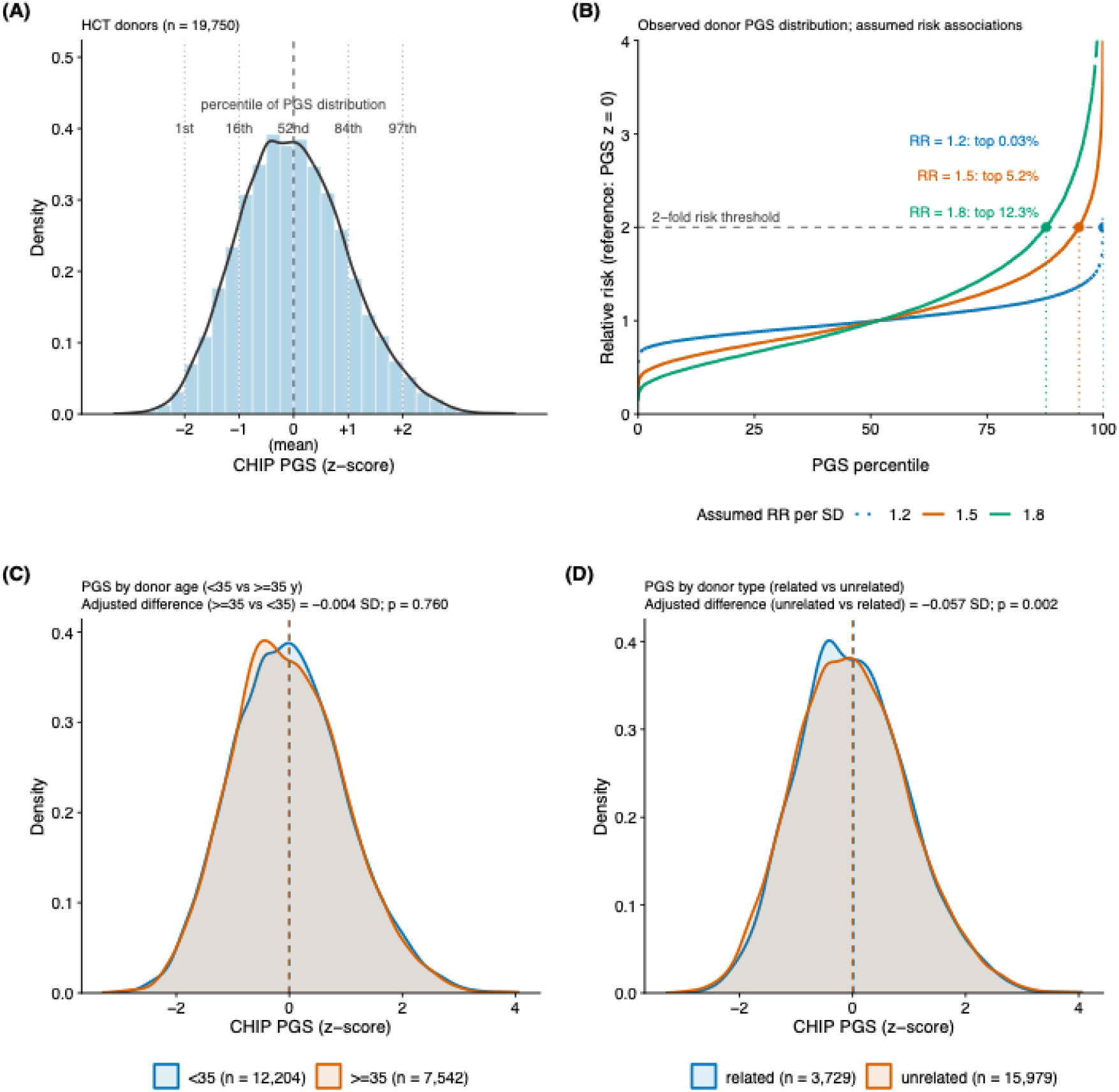
Inherited susceptibility to clonal hematopoiesis of indeterminate potential (CHIP) among HCT donors. (A) Distribution of standardized CHIP polygenic scores (PGSs) among 19,750 donors. Dotted vertical lines mark −2, −1, +1, and +2 standard deviations (SD); the dashed line marks the cohort mean (z-score = 0). Labels above each line give the corresponding percentile of the observed PGS distribution. (B) Illustration of how donor PGSs could translate into differences in risk of a hypothetical adverse outcome, assuming that each SD increase in PGS is associated with 20%, 50%, or 80% higher risk (relative risk [RR] of 1.2, 1.5, and 1.8). Risk is expressed relative to a donor with the average PGS (z-score = 0). The horizontal dashed line marks twofold risk; vertical dotted lines and points mark the PGS percentile at which each assumption reaches this threshold. Colored labels indicate the percentage of donors whose risk would reach or exceed twofold under each assumption. (C) Distribution of standardized CHIP PGSs among donors younger or older than 35 years. (D) Distribution of standardized CHIP PGSs among related versus unrelated donors. In (C) and (D), dashed vertical lines mark the overall mean, and the annotation reports the difference in mean PGS (in SD units) with its p-value, estimated from a linear model adjusted for the top eight genetic principal components.

Under a hypothetical assumption of a 20% increased risk of an adverse outcome (relative risk, RR = 1.2 per PGS SD), 0.03% of donors (5 of 19,750) would have at least twice the risk of a donor with the average PGS. This proportion increases substantially as the assumed risk per SD increases: under a 50% (RR = 1.5 per SD) and an 80% (RR = 1.8 per SD) risk model, the 5.2% and 12.3% of donors with highest PGS, respectively, would reach this twofold threshold (Figure 3B).

Using cohort-wide standardized PGS, no statistically significant differences in PGS distribution were noted between younger and older donors (<35 vs ≥35 years: adjusted difference −0.004 SD, p = 0.76) (Figure 3C). On the other hand, unrelated donors had a modestly but statistically significant lower adjusted mean PGS than related donors (adjusted difference −0.06 SD, p = 0.002; Figure 3D).

## Discussion

GS-BMT, the largest genomic investigation in HCT to date, holds great promise in refining donor selection, defining HCT outcomes, and advancing precision medicine for this population. In this manuscript, we showed that clinical factors alone have modest ability to predict survival after HCT, and that the use of different machine-learning algorithms was not superior to conventional time-to-event analysis. We also characterized inherited susceptibility to CHIP among donors, which was similar in younger and older donors, and illustrated how this could translate into differences in outcome risk. These findings highlight the importance of integrating genomic data with clinical information to improve risk prediction, optimize donor selection, and advance precision HCT practices.

Advances in machine learning (ML) algorithms enable the handling of large high dimensional clinical and multi-omic data to guide precision medicine approaches. Although ML in HCT is primarily limited to registry-based clinical information, several web-based tools have been developed such as the Sickle Cell Predicting Outcomes of Hematopoietic Cell Transplantation (SPRIGHT), and Myelofibrosis AllRisk stratifier.^16,17^ Yet, a systematic review of 27 studies that applied ML in HCT between 2015-2020 found insufficient evidence to determine the best ML technique or identify the minimal variables required in HCT setting.^18^ Consistent with this, our analyses showed that the more flexible ML approaches did not outperform the conventional Cox proportional hazards model for predicting post-HCT overall survival in AML using the available clinical variables. Specifically, the Cox model, RSF, and survival DNN demonstrated nearly identical discrimination and calibration. This finding is consistent with a growing body of comparative studies showing that ML methods often provide little or no improvement over conventional regression-based approaches for clinical prediction.^19–22^ Yet, the application of ML in GS-BMT with both clinical and genomic predictors may translate into improved performance because of its advantage of use in high-dimensional and correlated data such as genotyping..

Several recent studies highlighted the importance of germline genetic testing before HCT to guide patient risk stratification. Examples include testing for rare pathogenic germline variants in inherited bone marrow failure genes for patients with severe aplastic anemia or MDS, or DNA repair genes before radiation-based conditioning regimens.^23–25^ However, genetic screening of HCT donors remains focused on HLA compatibility and targeted testing based on the recipient’s medical history with some assessment of recipient post-HCT variants, usually CHIP, suspected to be of donor origin.^26^ Previous studies showed that the presence of CHIP variants in donors increased the risk of chronic G*v*HD and possibly donor-derived leukemia in HCT recipients.^27,28^ In our proof-of-concept analysis characterizing CHIP predisposition in HCT donors using polygenic genetic scores (PGS), we showed no differences in CHIP PGS by age consistent with their germline nature. Yet, CHIP PGS were higher in donors who were relatives of HCT recipients with hematological malignancies than unrelated donors. This is in line with reported associations from Mendelian randomization studies between CHIP PGSs and risk of CHIP mutations (OR per PGS-SD = 1.23) or certain hematological or solid tumors malignancies.^29–31^

Our illustrative analysis on donor CHIP PGS distribution showed it translates into differences in risk and therefore may be of use in donor selection. Genomic and clinical data from the GS-BMT will enable further investigation of whether donor inherited susceptibility to CHIP is associated with recipient outcomes, or age-related complications after HCT. This is particularly important in light of prior evidence of accelerated biological aging in HCT recipients.^32,33^

In conclusion, GS-BMT is a large registry-based multi-center study designed to establish the foundation for HCT genomics, where donor selection can be guided by key genomic markers, and patient treatment regimens can be personalized to their genetic background.

## Data Availability

The data produced in this study are under the authority of the Center for International Blood and Marrow Transplant Research (CIBMTR). Investigators may access these data from the CIBMTR through existing data access procedures.

## Acknowledgements

The study is funded, in part, by the intramural program of the National Cancer Institute (NCI), NIH. The CIBMTR is supported primarily by the Public Health Service U24CA076518 from the NCI, the National Heart, Lung and Blood Institute (NHLBI), and the National Institute of Allergy and Infectious Diseases (NIAID); 75R60222C00011 from the Health Resources and Services Administration (HRSA); and N00014-23-1-2057, N00014-24-1-2057, and N00014-25-214 from the Office of Naval Research. The authors acknowledge the research contributions of the Cancer Genomics Research Laboratory for their expertise, execution, and support of this research in the areas of project planning, wet laboratory processing of specimens, and bioinformatics analysis of generated data. This project has been funded in part with Federal funds from the National Cancer Institute, National Institutes of Health, under NCI Contract No. 75N910D00024. The contributions of the NIH author(s) were made as part of their official duties as NIH federal employees, are in compliance with agency policy requirements, and are considered Works of the United States Government. The findings and conclusions presented in this paper are those of the author(s) and do not necessarily reflect the views or policies of the NIH or the U.S. Department of Health and Human Services, nor does mention of trade names, commercial products, or organizations imply endorsement by the U.S. Government.

## References

1. Spellman SR, Xu K, Oloyede T, et al. Current Activity Trends and Outcomes in Hematopoietic Cell Transplantation and Cellular Therapy - A Report from the CIBMTR. Transplant Cell Ther. Aug 2025;31(8):505–532. doi:10.1016/j.jtct.2025.05.014

2. D’Souza A, Lee S, Zhu X, Pasquini M. Current Use and Trends in Hematopoietic Cell Transplantation in the United States. Biol Blood Marrow Transplant. Sep 2017;23(9):1417–1421. doi:10.1016/j.bbmt.2017.05.035

3. Current Uses and Outcomes of Cellular Therapy in the US, Summary Slides 2025.

4. Sanz J, Galimard JE, Labopin M, et al. Post-transplant cyclophosphamide after matched sibling, unrelated and haploidentical donor transplants in patients with acute myeloid leukemia: a comparative study of the ALWP EBMT. J Hematol Oncol. May 6 2020;13(1):46. doi:10.1186/s13045-020-00882-6

5. Zeiser R, von Bubnoff N, Butler J, et al. Ruxolitinib for Glucocorticoid-Refractory Acute Graft-versus-Host Disease. N Engl J Med. May 7 2020;382(19):1800–1810. doi:10.1056/NEJMoa1917635

6. Beelen DW, Stelljes M, Remenyi P, et al. Treosulfan compared with reduced-intensity busulfan improves allogeneic hematopoietic cell transplantation outcomes of older acute myeloid leukemia and myelodysplastic syndrome patients: Final analysis of a prospective randomized trial. Am J Hematol. Aug 2022;97(8):1023–1034. doi:10.1002/ajh.26620

7. Karaesmen E, Rizvi AA, Preus LM, et al. Replication and validation of genetic polymorphisms associated with survival after allogeneic blood or marrow transplant. Blood. Sep 28 2017;130(13):1585–1596. doi:10.1182/blood-2017-05-784637

8. Savage SA, Chanock S. Genetic variation and hematopoietic stem cell transplantation: expansion of the paradigm. Pediatr Transplant. 2003;7 Suppl 3:32–9.

9. Sucheston-Campbell LE, Clay A, McCarthy PL, et al. Identification and utilization of donor and recipient genetic variants to predict survival after HCT: are we ready for primetime? Curr Hematol Malig Rep. Mar 2015;10(1):45–58. doi:10.1007/s11899-014-0246-x

10. Perez-Garcia A, De la Camara R, Roman-Gomez J, et al. CTLA-4 polymorphisms and clinical outcome after allogeneic stem cell transplantation from HLA-identical sibling donors. Blood. Jul 1 2007;110(1):461–7. doi:10.1182/blood-2007-01-069781

11. Jagasia M, Clark WB, Brown-Gentry KD, et al. Genetic variation in donor CTLA-4 regulatory region is a strong predictor of outcome after allogeneic hematopoietic cell transplantation for hematologic malignancies. Biol Blood Marrow Transplant. Jul 2012;18(7):1069–75. doi:10.1016/j.bbmt.2011.12.518

12. Gadalla SM, Wang Y, Wang T, et al. Association of donor IFNL4 genotype and non-relapse mortality after unrelated donor myeloablative haematopoietic stem-cell transplantation for acute leukaemia: a retrospective cohort study. Lancet Haematol. Oct 2020;7(10):e715–e723. doi:10.1016/S2352-3026(20)30294-5

13. Hahn T, Wang J, Preus LM, et al. Novel genetic variants associated with mortality after unrelated donor allogeneic hematopoietic cell transplantation. EClinicalMedicine. Oct 2021;40:101093. doi:10.1016/j.eclinm.2021.101093

14. Zhu Q, Yan L, Liu Q, et al. Exome chip analyses identify genes affecting mortality after HLA-matched unrelated-donor blood and marrow transplantation. Blood. May 31 2018;131(22):2490–2499. doi:10.1182/blood-2017-11-817973

15. Kessler MD, Damask A, O’Keeffe S, et al. Common and rare variant associations with clonal haematopoiesis phenotypes. Nature. Dec 2022;612(7939):301–309. doi:10.1038/s41586-022-05448-9

16. Hernandez-Boluda JC, Mosquera-Orgueira A, Gras L, et al. Use of machine learning techniques to predict poor survival after hematopoietic cell transplantation for myelofibrosis. Blood. Jun 26 2025;145(26):3139–3152. doi:10.1182/blood.2024027287

17. Chandrasekar RS, Kane M, Krishnamurti L. Machine-Learning Predictive Tool for the Individualized Prediction of Outcomes of Hematopoietic Cell Transplantation for Sickle Cell Disease: Registry-Based Study. JMIR AI. Sep 15 2025;4:e64519. doi:10.2196/64519

18. Gupta V, Braun TM, Chowdhury M, Tewari M, Choi SW. A Systematic Review of Machine Learning Techniques in Hematopoietic Stem Cell Transplantation (HSCT). Sensors (Basel). Oct 27 2020;20(21)doi:10.3390/s20216100

19. Hu Y, Zhang X, Slavin V, et al. Beyond Comparing Machine Learning and Logistic Regression in Clinical Prediction Modelling: Shifting from Model Debate to Data Quality. J Med Internet Res. Nov 5 2025;27:e77721. doi:10.2196/77721

20. Baralou V, Kalpourtzi N, Touloumi G. Individual risk prediction: Comparing random forests with Cox proportional-hazards model by a simulation study. Biom J. Aug 2023;65(6):e2100380. doi:10.1002/bimj.202100380

21. Christodoulou E, Ma J, Collins GS, Steyerberg EW, Verbakel JY, Van Calster B. A systematic review shows no performance benefit of machine learning over logistic regression for clinical prediction models. J Clin Epidemiol. Jun 2019;110:12–22. doi:10.1016/j.jclinepi.2019.02.004

22. Birolo G, Rossi I, Sartori F, Rollo C, Fariselli P, Sanavia T. Beyond Cox models: Assessing the performance of machine-learning methods in non-proportional hazards and non-linear survival analysis. Comput Biol Med. Nov 2025;198(Pt B):111176. doi:10.1016/j.compbiomed.2025.111176

23. McReynolds LJ, Rafati M, Wang Y, et al. Genetic testing in severe aplastic anemia is required for optimal hematopoietic cell transplant outcomes. Blood. Aug 25 2022;140(8):909–921. doi:10.1182/blood.2022016508

24. Feurstein S, Trottier AM, Estrada-Merly N, et al. Germ line predisposition variants occur in myelodysplastic syndrome patients of all ages. Blood. Dec 15 2022;140(24):2533–2548. doi:10.1182/blood.2022015790

25. Rafati M, Wang Y, Koppayi AL, et al. Germline Pathogenic Variants in MUTYH Are Associated With Inferior Survival After Hematopoietic Cell Transplantation in Patients With Hematologic Malignancies or Disorders. Am J Hematol. Dec 2025;100(12):2442–2447. doi:10.1002/ajh.70093

26. Pryce A, Van Eerden E, Cody M, et al. Genetic Findings of Potential Donor Origin following Hematopoietic Cell Transplantation: Recommendations on Donor Disclosure and Genetic Testing from the World Marrow Donor Association. Transplant Cell Ther. Feb 2024;30(2):143–154. doi:10.1016/j.jtct.2023.11.019

27. Rojek K, Nickels E, Neistadt B, et al. Identifying Inherited and Acquired Genetic Factors Involved in Poor Stem Cell Mobilization and Donor-Derived Malignancy. Biol Blood Marrow Transplant. Nov 2016;22(11):2100–2103. doi:10.1016/j.bbmt.2016.08.002

28. Newell LF, Williams T, Liu J, et al. Engrafted Donor-Derived Clonal Hematopoiesis after Allogenic Hematopoietic Cell Transplantation is Associated with Chronic Graft-versus-Host Disease Requiring Immunosuppressive Therapy, but no Adverse Impact on Overall Survival or Relapse. Transplant Cell Ther. Aug 2021;27(8):662 e1–662 e9. doi:10.1016/j.jtct.2021.04.014

29. Uddin MM, Saadatagah S, Niroula A, et al. Long-term longitudinal analysis of 4,187 participants reveals insights into determinants of clonal hematopoiesis. Nat Commun. Sep 9 2024;15(1):7858. doi:10.1038/s41467-024-52302-9

30. Kar SP, Quiros PM, Gu M, et al. Genome-wide analyses of 200,453 individuals yield new insights into the causes and consequences of clonal hematopoiesis. Nat Genet. Aug 2022;54(8):1155–1166. doi:10.1038/s41588-022-01121-z

31. Ranasinghe D, Lin WY, Fordham SE, et al. Common variation at 1q23.3, 2p23.3, 2q33.3, and 2p21 influences the risk of acute myeloid leukemia. Blood. Apr 23 2026;147(17):1958–1969. doi:10.1182/blood.2025031266

32. Alsaggaf R, Katta S, Wang T, et al. Epigenetic Aging and Hematopoietic Cell Transplantation in Patients With Severe Aplastic Anemia. Transplant Cell Ther. Apr 2021;27(4):313 e1–313 e8. doi:10.1016/j.jtct.2021.01.013

33. Gadalla SM, Katki HA, Lai TP, et al. Donor telomeres and their magnitude of shortening post-allogeneic haematopoietic cell transplant impact survival for patients with early-stage leukaemia or myelodysplastic syndrome. EBioMedicine. Apr 2025;114:105641. doi:10.1016/j.ebiom.2025.105641

