## Supplemental Materials for "Genomic Studies in Blood and Marrow Transplantation (GS-BMT): An NCI-CIBMTR genomic medicine initiative in hematopoietic cell transplantation"

**Supplemental Methods**

*Genotyping, Imputation and Quality Control*

Variant- and sample-level genotyping quality control (QC) was performed using a QC pipeline developed by the NCI Division of Cancer Epidemiology and Genetics Cancer Genomics Research (CGR) Laboratory. Genotypes were called using the standard Illumina array-analysis workflow with a study-specific cluster file. A two-stage completion rate filter was applied, with locus filters followed by sample filters at each stage. This used an initial threshold of 0.80 for both loci and samples, followed by a subsequent threshold of 0.95 for both loci and samples. Sample contamination was estimated using VerifyIDintensity with 1000 Genomes B-allele frequencies as the reference, and samples with predicted contamination greater than 10% were excluded. Samples failing initial QC filters were subject to recovery efforts where possible, including STR fingerprinting (Identifiler) to confirm sample identity and screen for contamination.

Reported sex was verified against genotypically inferred sex based on the X chromosome method-of-moments F coefficient calculated in PLINK, with an assignment threshold of 0.5 (midpoint between the expected values of 0 for females and 1 for males). Samples with discordant genetic and reported sex were subjected to additional screening for sex chromosome aneuploidies (e.g., XXY and X0). Chromosome X and Y copy-number variation data were reviewed, and the reported sex was confirmed with the investigator before any samples with unresolved sex discordance were excluded from the final imputed dataset.

Genetic ancestry was inferred with GRAF-pop, which projects each subject into a low dimensional space based on their similarity to a set of African, Asian, European, South Asian, and Latin American reference samples drawn from several dbGAP projects, with sample labels assigned based on study-reported population values. Genetic ancestry groups are then defined using cutoffs chosen by GRAF-pop developers to group an independent set of dbGAP samples based on study-reported population labels. These genetic ancestry groups are specific to these cutoffs and reflect the reference samples and choices made by GRAF-pop developers; they are presented here for quality control and for comparison purposes. Autosomal heterozygosity was assessed within each ancestry group (limited to groups with more than 50 subjects) using the PLINK method-of-moments F coefficient, with subjects falling outside ±0.20 flagged as potential outliers and excluded from the final imputed dataset. Pairwise relatedness was estimated separately within each ancestry group using PLINK identity-by-descent (IBD) analysis on LD-pruned markers, with PI_HAT greater than 0.2 defining first- or second-degree relatedness. Genetic principal components analysis (PCA) was performed using smartpca (EIGENSOFT) within each genetically inferred ancestry group on a set of minor allele frequency (MAF)- and LD-pruned markers.

For imputation, GS-BMT genotyped samples passing QC were merged with those from 84,731 participants in the NCI Prostate, Lung, Colorectal, and Ovarian (PLCO) Cancer Screening Trial, which serves as an independent comparison group for downstream disease etiology analyses. PLCO samples were genotyped on a comparable Illumina Global Screening Array and were lifted over from GRCh37 to GRCh38. SNP outliers driven by array or batch differences between the two datasets were identified and removed through allele frequency comparison and association testing between datasets. The combined dataset was then prepared for imputation against the TOPMed r3 reference panel using the TOPMed Imputation Server. Variants were filtered for departure from Hardy-Weinberg equilibrium tested separately within each ancestry group in unrelated subjects (autosomal SNPs with MAF > 0.01; P < 1×10⁻⁶). Additional pre-imputation QC was performed using the Will Rayner conversion toolkit (https://www.chg.ox.ac.uk/~wrayner/tools/), which enforces compatibility with the reference panel by aligning strand orientation and allele assignments, and removing palindromic variants with minor allele frequency > 0.4, variants with allele frequency differences greater than 0.2 versus the reference, variants with alleles differing from or absent from the reference, indels, and duplicates. Variants with imputation quality Rsq < 0.2 were excluded from the final dataset, which was converted to PGEN format using PLINK2 for downstream analyses.

*Prediction Tools for Clinical Factors Impacting HCT Survival for patients with AML*

We evaluated three complementary approaches for predicting post-HCT overall survival: a Cox proportional hazards model, a random survival forest (RSF), and a deep neural network survival model (survival DNN). The multivariable Cox proportional hazards model^1^ was fitted using all 15 covariates. Hazard ratios (HRs), 95% confidence intervals (CIs), and Wald *p*-values were estimated. Individual risk scores were derived from the linear predictor. Proportional hazards assumption was tested using the Schoenfeld residuals test (Table S2).

The RSF is a nonparametric survival modeling approach that does not assume linear effects or proportional hazards and can capture nonlinear associations and higher-order interactions through recursive tree splitting^2^. In this study, RSF was trained using 500 trees with a minimum node size of 15.  For each tree, splitting was based on log-rank statistics. RSF estimates risk by aggregating cumulative hazard estimates across an ensemble of survival trees. For RSF sensitivity analyses, we compared models using cross-validation-selected tuning parameters with models using default settings. The predictive performance of cross-validation, assessed using the test C-index and integrated Brier score, was highly similar between the tuned and default models, with negligible differences in predictive performance. Therefore, we used the default RSF tuning parameters in the primary analysis. Variable importance was assessed using permutation-based importance, defined as the decrease in C-index following random permutation of each predictor.

For survival DNN, we fit a feed-forward neural network adapted for right-censored survival data by training on the Cox partial likelihood^3^. This approach allows the model to capture nonlinear associations and interactions among predictors while accounting for censoring. Compared with a standard Cox model, the survival DNN replaces the linear predictor with a neural network-based risk score. Hyperparameters, including the number and size of hidden layers, learning rate, regularization, and number of training epochs, were selected using 5-fold cross-validation within the training data. The configuration with the highest mean concordance index was retained. In the full analysis, the selected configuration included two hidden layers with 32 and 16 units, a learning rate of 0.001, Cox partial likelihood loss with L2 regularization, and 200 training epochs. Because the model was trained using the Cox partial likelihood, it follows the proportional hazards framework while allowing nonlinear relationship between predictors and the risk score. Variable importance was assessed by permutation importance, defined as the decrease in C-index after shuffling each predictor.

For each of the fitted survival model, we calculated a continuous, model-specific risk score for every patient. The risk score represents the patient’s predicted relative risk of death, with higher values indicating higher predicted mortality risk. We then divided patients into three risk groups. Patients with risk scores below the 33.3^rd^ percentile were classified as the **low-risk group**, those between the 33.3^rd^ and 66.7^th^ percentiles were classified as the **intermediate-risk group**, and those above the 66.7^th^ percentile were classified as the **high-risk group**. Kaplan–Meier curves were used to evaluate survival differences across groups, with statistical comparison via the log-rank test. Agreement in risk classification for the high-risk and low-risk group was assessed by comparing overlap across models, visualized using an UpSet plot. Predictive performance was evaluated using 100 repeated random train-test splits, with 80% of patients assigned to the training set and 20% to the testing set, stratified by AML stage and vital status. We calculated C-index for model discrimination (ability to correctly rank patients according to risk) and integrated Brier score (IBS) for prediction errors over the follow-up period^4^.

Statistical analyses were performed using R (version 4.5.2). Cox proportional hazards models and Kaplan-Meier analyses were fitted using the survival package. Random survival forests were fitted using randomForestSRC, survival DNNs using survdnn, prediction error curves and integrated Brier scores using pec, and figures were generated using ggplot2.

| **Table S1**. Index variants and weights used to construct the germline polygenic score for clonal hematopoiesis of indeterminate potential. | | | | | | | |
| --- | --- | --- | --- | --- | --- | --- | --- |
| **rsID** | **Chromosome** | **Position (build 38)** | **Other allele** | **Effect allele** | **Effect allele frequency** | **Effect allele Beta** | ***P*** |
| rs80019658 | 11 | 108043157 | T | C | 0.13 | -0.08 | 9.92E-09 |
| rs10890839 | 11 | 108435509 | C | A | 0.41 | 0.11 | 1.43E-32 |
| rs149934734 | 11 | 108444879 | C | T | 0.03 | 0.21 | 3.97E-13 |
| rs16930705 | 12 | 26436837 | G | A | 0.07 | 0.17 | 3.72E-22 |
| rs72698720 | 14 | 100712378 | C | G | 0.14 | -0.08 | 5.08E-09 |
| rs78378222 | 17 | 7668434 | T | G | 0.01 | 0.26 | 1.64E-09 |
| rs7406485 | 17 | 7827723 | C | G | 0.32 | -0.07 | 1.67E-11 |
| rs80093687 | 17 | 57386496 | C | T | 0.05 | -0.18 | 3.55E-16 |
| rs8088824 | 18 | 44571296 | C | T | 0.76 | -0.08 | 3.57E-14 |
| rs62175229 | 2 | 159821048 | C | T | 0.56 | 0.07 | 5.6E-13 |
| rs78446341 | 2 | 159834145 | G | A | 0.02 | -0.18 | 1.39E-08 |
| rs12472767 | 2 | 159925824 | T | C | 0.23 | -0.07 | 3.02E-11 |
| rs7272062 | 20 | 31671342 | A | G | 0.21 | -0.08 | 6.07E-11 |
| rs2834708 | 21 | 34972056 | C | T | 0.36 | 0.06 | 1.66E-10 |
| rs62235635 | 22 | 27918624 | G | A | 0.01 | 0.30 | 4.76E-08 |
| rs361725 | 22 | 50532618 | T | C | 0.63 | -0.05 | 2.47E-08 |
| rs6440667 | 3 | 150287063 | C | G | 0.82 | -0.08 | 6.72E-10 |
| rs56139929 | 3 | 160106004 | C | T | 0.11 | 0.10 | 6.79E-12 |
| rs2305407 | 3 | 160412322 | G | A | 0.53 | 0.12 | 3.63E-36 |
| rs75008921 | 3 | 160440785 | C | T | 0.08 | -0.10 | 5.69E-09 |
| rs79683816 | 3 | 160558751 | CT | C | 0.01 | 0.40 | 1.67E-22 |
| rs144317085 | 4 | 104884951 | A | T | 0.03 | 0.19 | 1.03E-14 |
| rs4975592 | 5 | 1097331 | G | A | 0.42 | -0.06 | 1.93E-11 |
| rs71593392 | 5 | 1203384 | T | C | 0.29 | -0.06 | 1.36E-08 |
| rs59438556 | 5 | 1228120 | CT | C | 0.61 | 0.06 | 3.7E-09 |
| rs112333466 | 5 | 1249701 | C | T | 0.01 | 0.24 | 2.58E-10 |
| rs2853690 | 5 | 1253629 | G | A | 0.19 | -0.09 | 4.73E-13 |
| rs35812074 | 5 | 1267766 | C | G | 0.02 | 0.29 | 1.36E-18 |
| rs2075785 | 5 | 1278469 | C | T | 0.12 | -0.12 | 2.71E-16 |
| rs13167280 | 5 | 1280362 | G | A | 0.13 | 0.15 | 4.53E-29 |
| rs7705526 | 5 | 1285859 | C | A | 0.32 | 0.25 | 1.29E-140 |
| rs112290073 | 5 | 1285917 | G | A | 0.01 | 0.24 | 5.43E-09 |
| rs2853676 | 5 | 1288432 | T | C | 0.73 | -0.13 | 1.14E-34 |
| rs2736098 | 5 | 1293971 | C | T | 0.28 | 0.15 | 1.82E-50 |
| rs33987166 | 5 | 1296643 | T | C | 0.03 | 0.21 | 7.43E-14 |
| rs4975612 | 5 | 1300195 | G | T | 0.22 | -0.11 | 1.06E-22 |
| rs543796723 | 5 | 1303752 | CT | C | 0.01 | 0.38 | 2.02E-11 |
| rs35953391 | 5 | 1312214 | C | T | 0.20 | -0.14 | 5.52E-34 |
| rs149462697 | 5 | 1361187 | G | C | 0.02 | -0.23 | 3.57E-08 |
| rs27919 | 5 | 1361773 | T | C | 0.89 | -0.12 | 9.11E-15 |
| rs62329730 | 5 | 1366259 | G | A | 0.02 | 0.20 | 1.05E-10 |
| rs2086132 | 5 | 1372402 | G | C | 0.27 | 0.09 | 2.77E-16 |
| rs11564750 | 5 | 1447647 | G | C | 0.10 | -0.09 | 3.6E-08 |
| rs2396004 | 6 | 43388113 | A | G | 0.57 | -0.05 | 1.84E-08 |
| rs4945828 | 6 | 109262379 | C | T | 0.80 | -0.08 | 1.45E-11 |
| rs2356817 | 6 | 109321472 | G | A | 0.48 | -0.11 | 1.38E-29 |
| rs2275652 | 6 | 109382480 | G | C | 0.30 | -0.13 | 1.39E-34 |
| rs752261 | 6 | 109449666 | G | A | 0.06 | 0.11 | 3.46E-09 |
| rs145655884 | 6 | 109499851 | C | T | 0.04 | -0.16 | 5.72E-10 |
| rs11762008 | 7 | 44864065 | A | G | 0.43 | 0.07 | 4.5E-14 |
| rs7817503 | 8 | 129563195 | T | C | 0.51 | 0.06 | 5.66E-10 |

**Table S2**: Multivariable Cox proportional hazards model for post-HCT mortality

| **Variable** | **Hazard Ratio (95% CI)** | ***P*** |
| --- | --- | --- |
| **AML stage at HCT** |  |  |
| CR1 (1^st^ complete remission) | Reference |  |
| CR2 | 1.10 (1.02 – 1.19) | .013 |
| CR3+ | 1.45 (1.14 – 1.84) | .002 |
| Active disease | 1.93 (1.81 – 2.06) | <.001 |
| **Donor type** |  |  |
| HLA-identical sibling | Reference |  |
| Well-matched URD (8/8) | 1.26 (1.14 – 1.40) | <.001 |
| Partially-matched URD (7/8) | 1.67 (1.48 – 1.89) | <.001 |
| Mis-matched URD (6/8) | 2.54 (1.86 – 3.46) | <.001 |
| Haploidentical donor | 1.36 (1.16 – 1.60) | <.001 |
| **HCT-comorbidity index** |  |  |
| 0 | Reference |  |
| 1 | 1.05 (0.96 – 1.15) | .293 |
| 2 | 1.13 (1.03 – 1.24) | .009 |
| 3+ | 1.35 (1.26 – 1.46) | <.001 |
| **Karnofsky score** |  |  |
| < 90 | Reference |  |
| 90-100 | 0.91 (0.86 – 0.96) | <.001 |
| **Stem cell Source** |  |  |
| Bone marrow | Reference |  |
| Peripheral blood | 1.01 (0.94 – 1.09) | .812 |
| **Recipient age (per year)** | 1.01 (1.01 – 1.01) | <.001 |
| **Donor age (per year)** | 1.01 (1.00 – 1.01) | <.001 |
| **Conditioning regimen intensity** |  |  |
| Myeloablative | Reference |  |
| Non-myeloablative/reduced intensity | 1.06 (0.98 – 1.14) | .128 |
| **Total body irradiation-regimen** |  |  |
| No | Reference |  |
| Yes | 1.13 (1.06 – 1.22) | <.001 |
| **GvHD prophylaxis** |  |  |
| CSA | Reference |  |
| Ex vivo TCD/CD34 | 0.70 (0.57 – 0.87) | <.001 |
| Other prophylaxis | 0.65 (0.49 – 0.86) | .003 |
| PTCy+/-others | 0.68 (0.59 – 0.79) | <.001 |
| Tac+/-others | 0.82 (0.75 – 0.90) | <.001 |
| **ATG or Campath use** |  |  |
| None | Reference |  |
| ATG | 0.98 (0.92 – 1.04) | .520 |
| Campath | 1.04 (0.88 – 1.24) | .650 |
| **Recipient sex** |  |  |
| Female | Reference |  |
| Male | 1.06 (1.00 – 1.12) | .040 |
| **Donor sex** |  |  |
| Female | Reference |  |
| Male | 0.94 (0.89 – 0.99) | .031 |
| **Donor-recipient CMV sero-status** |  |  |
| -/- | Reference |  |
| -/+ | 1.15 (1.07 – 1.23) | <.001 |
| +/- | 0.97 (0.86 – 1.09) | .612 |
| +/+ | 1.06 (0.98 – 1.15) | .143 |
| **Transplant year** |  |  |
| 2000 - 2009 | Reference |  |
| 2010 - 2014 | 0.90 (0.82 – 0.99) | .026 |
| 2015 - 2019 | 0.73 (0.66 – 0.81) | <.001 |

Table S3. Schoenfeld residual test for proportional hazards

| **Variable** | $\boldsymbol{\chi}^{\boldsymbol{2}}$ | **df** | ***P*** |
| --- | --- | --- | --- |
| AML Stage at HCT | 24.774 | 3 | <.001 |
| Donor Type | 20.618 | 4 | <.001 |
| HCT-comorbidity index | 4.503 | 3 | .212 |
| Karnofsky Score | 5.565 | 1 | .018 |
| Stem Cell Source | 25.583 | 1 | <.001 |
| Recipient age | 30.585 | 1 | <.001 |
| Donor age | 0.441 | 1 | .507 |
| Conditioning regimen intensity | 39.482 | 1 | <.001 |
| TBI-regimens | 4.937 | 1 | .026 |
| GvHD prophylaxis | 7.148 | 4 | .128 |
| ATG or Campath use | 15.065 | 2 | <.001 |
| Recipient sex | 13.702 | 1 | <.001 |
| Donor sex | 0.621 | 1 | .431 |
| Donor-recipient CMV sero-status | 0.233 | 3 | .972 |
| Transplant year | 45.000 | 2 | <.001 |
| GLOBAL | 175.654 | 29 | <.001 |

Table S4. Spearman Correlation of rank of model-specific risk scores.

| **Models** | **Spearman Correlation** |
| --- | --- |
| RSF *vs* Cox | 0.898 |
| Survival DNN *vs* Cox | 0.964 |
| Survival DNN *vs* RSF | 0.906 |
| Abbreviations, RSF: random survival forest; Cox: Cox proportional hazards model; Survival DNN: deep neural network survival model. | |

**Figure S1**: Overlap in patient risk classification among the three models.

UpSet plots show the number of patients in each possible intersection of risk group classification assigned by the Cox proportional hazards model (Cox), random survival forest (RSF), and survival deep neural network (SurvDNN). (A) high-risk and (B) low-risk groups. Each column represents a unique intersection pattern indicated by the connected dots.

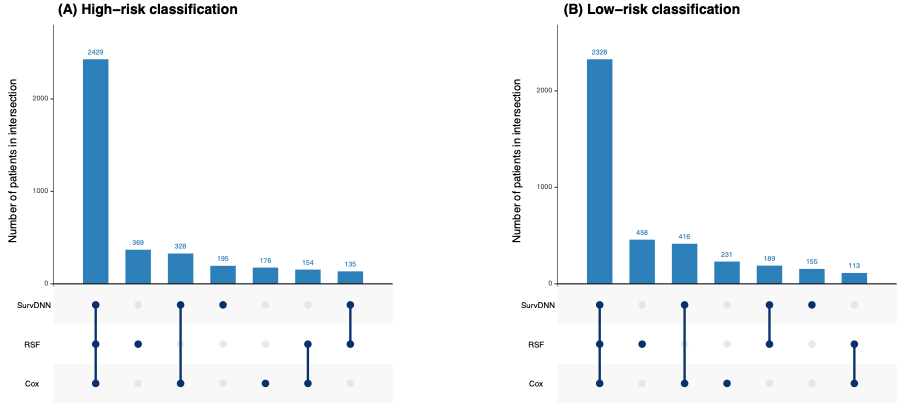

**Figure S2:** Time-dependent Brier score curves RSF, Survival DNN, and Cox proportional hazards models. Reference indicates a Kaplan–Meier null model

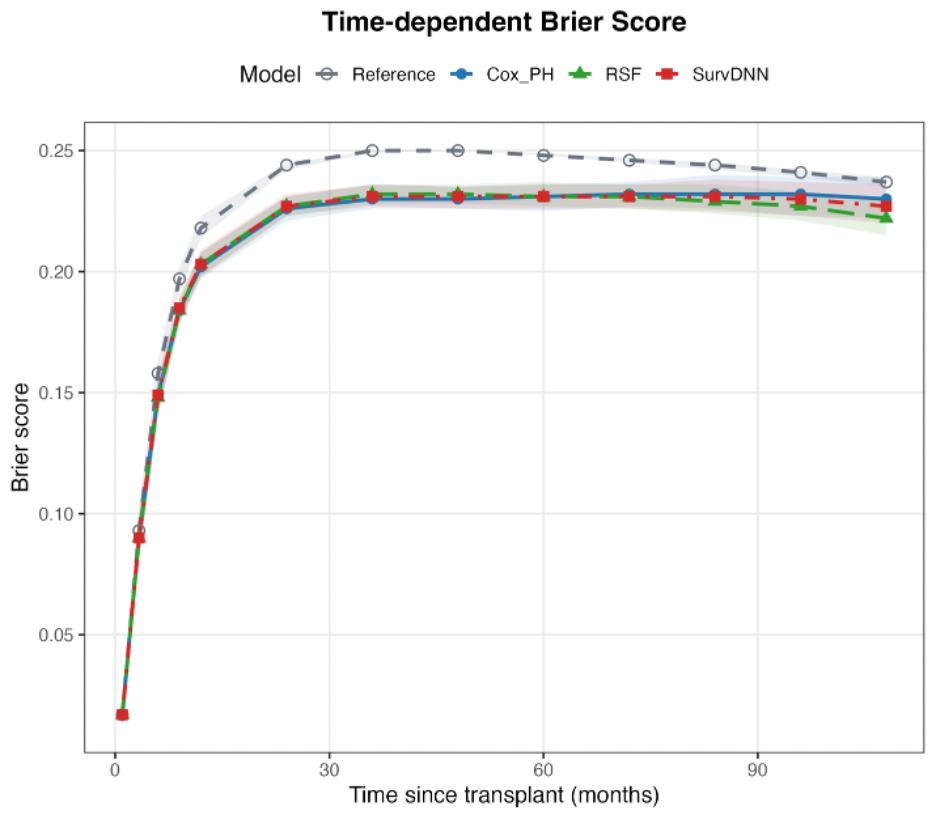

**Figure S3:** Subject genetic distance as a proportion of the ancestral population. Each subject is projected as an admixed proportion of three base ancestral populations (African, Asian, and European)

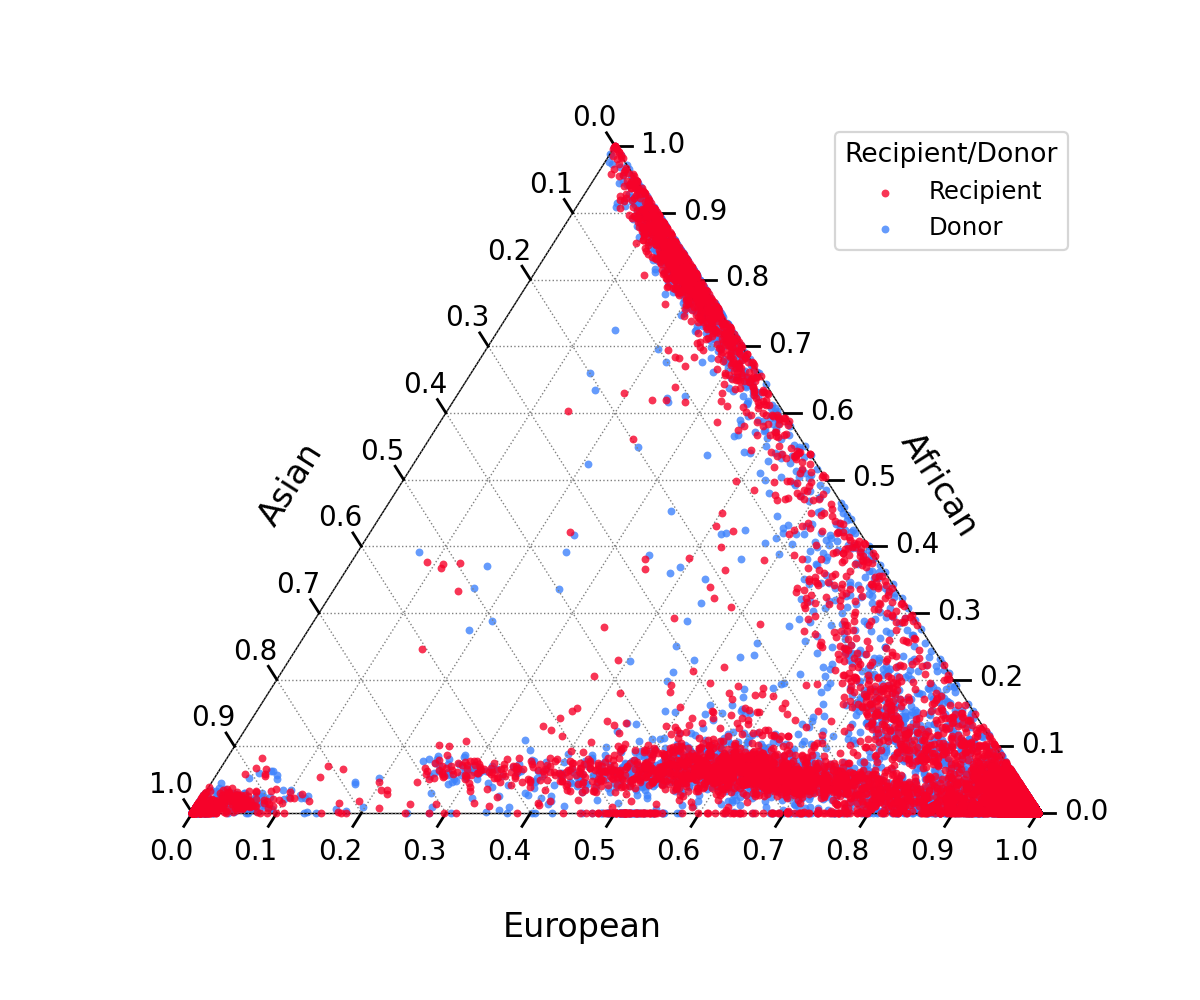

**References**

1.	Cox D. Regression models and life‐tables. *Journal of the royal statistical society: Series B (methodological)*. 1972;34(2):187-202.

2.	Ishwaran H, Kogalur U, Blackstone E, MS. L. Random survival forests. *The Annals of Applied Statistics, Ann Appl Stat* 2008;2(3):841-860.

3.	Kvamme H, Borgan Ø, Scheel I. Time-to-event prediction with neural networks and Cox regression. *Journal of machine learning research*. 2019;20(129):1-30.

4.	Graf E, Schmoor C, Sauerbrei W, Schumacher M. Assessment and comparison of prognostic classification schemes for survival data. *Stat Med*. Sep 15-30 1999;18(17-18):2529-45. doi:10.1002/(sici)1097-0258(19990915/30)18:17/18<2529::aid-sim274>3.0.co;2-5
